# Disparities in SARS-CoV-2 seroprevalence among individuals presenting for care in central North Carolina over a six-month period

**DOI:** 10.1101/2021.03.25.21254320

**Authors:** Cesar A. Lopez, Clark H. Cunningham, Sierra Pugh, Katerina Brandt, Usaphea P. Vanna, Matthew J. Delacruz, Quique Guerra, Samuel Jacob Goldstein, Yixuan J. Hou, Margaret Gearhart, Christine Wiethorn, Candace Pope, Carolyn Amditis, Kathryn Pruitt, Cinthia Newberry-Dillon, John Schmitz, Lakshmanane Premkumar, Adaora A. Adimora, Michael Emch, Ross Boyce, Allison E. Aiello, Bailey K. Fosdick, Daniel B. Larremore, Aravinda M. de Silva, Jonathan J Juliano, Alena J. Markmann

## Abstract

**Background:** Robust community-level SARS-CoV-2 prevalence estimates have been difficult to obtain in the American South and outside of major metropolitan areas. Furthermore, though some previous studies have investigated the association of demographic factors such as race with SARS-CoV-2 exposure risk, fewer have correlated exposure risk to surrogates for socioeconomic status such as health insurance coverage.

**Methods:** We used a highly specific serological assay utilizing the receptor binding domain of the SARS-CoV-2 spike-protein to identify SARS-CoV-2 antibodies in remnant blood samples collected by the University of North Carolina Health system. We estimated the prevalence of SARS-CoV-2 in this cohort with Bayesian regression, as well as the association of critical demographic factors with higher prevalence odds.

**Findings:** Between April 21^st^ and October 3^rd^ of 2020, a total of 9,624 unique samples were collected from clinical sites in central NC and we observed a seroprevalence increase from 2·9 (1·7, 4·3) to 9·1 (7·2, 11·1) over the study period. Individuals who identified as Latinx were associated with the highest odds ratio of SARS-CoV-2 exposure at 7·77 overall (5·20, 12·10). Increased odds were also observed among Black individuals and individuals without public or private health insurance.

**Interpretation:** Our data suggests that for this care-accessing cohort, SARS-CoV-2 seroprevalence was significantly higher than cumulative total cases reported for the study geographical area six months into the COVID-19 pandemic in North Carolina. The increased odds of seropositivity by ethnoracial grouping as well as health insurance highlights the urgent and ongoing need to address underlying health and social disparities in these populations.

**RESEARCH IN CONTEXT:** *Evidence before this study:* We searched PubMed for studies published through March 21^st^, 2021. We used search terms that included “COVID-19”, “SARS-CoV-2”, “prevalence” and “seroprevalence”. Our search resulted in 399 papers, from which we identified 58 relevant studies describing SARS-CoV-2 seroprevalence at sites around the United States from March 1 to December 9, 2020, 12 of which utilized remnant clinical samples and three of which overlapped with our study area. Most notably, one study of 4,422 asymptomatic inpatients and outpatients in central NC from April 28-June 19, 2020 found an estimated seroprevalence of 0·7 −0·8%, and another study of 177,919 inpatients and outpatients (3,817 from NC) from July 27-September 24, 2020 found an estimated seroprevalence of 2·5 −6·8%.

*Added value of this study:* This is the largest SARS-CoV-2 seroprevalence cohort published to date in NC. Importantly, we used a Bayesian framework to account for uncertainty in antibody assay sensitivity and specificity and investigated seropositivity by important demographic variables that have not yet been studied in this context in NC. This study corroborates other reports that specific demographic factors including race, ethnicity and the lack of public or private insurance are associated with elevated risk of SARS-CoV-2 infection. Furthermore, in a subset of serum samples, we identify other SARS-CoV-2 antibodies elicited by these individuals, including functionally neutralizing antibodies.

*Implications of all the available evidence:* It is difficult to say the exact seroprevalence in the central North Carolina area, but a greater proportion of the population accessing healthcare has been infected by SARS-CoV-2 than is reflected by infection cases confirmed by molecular testing. Furthermore, local governments need to prioritize addressing the many forms of systemic racism and socioeconomic disadvantage that drive SARS-CoV-2 exposure risk, such as residential and occupational risk, and an urgent need to provide access to SARS-CoV-2 testing and vaccination to these groups.

## INTRODUCTION

In December 2019, a cluster of pneumonia cases in China’s Hubei province heralded the beginning of what would become a global pandemic caused by severe acute respiratory syndrome coronavirus 2 (SARS-CoV-2). Despite attempts to contain the virus, SARS-CoV-2 has spread around the world, causing over 100 million infections and over 2 million deaths due to the respiratory disease it causes, COVID-19.^1^ Serological testing complements molecular testing for evaluating the spread of SARS-CoV-2 and can be deployed efficiently at the population level.^2^ Recently, large prevalence studies around the United States using remnant samples from healthcare settings have reported substantial geographic variation in prevalence by state: around 30% in New York but less than 2% in North Carolina (NC), the focus of the present study.^3,4^ Notably, two other studies overlap with the present cohort both temporally and geographically. One study of 4,422 asymptomatic inpatients and outpatients in central NC from April 28-June 19, 2020 found an estimated seroprevalence of 0 ·7 - 0·8%, and another study of 177,919 remnant clinical laboratory samples from routine screening (3,817 from NC) from July 27-September 24, 2020 found an estimated seroprevalence of 2·5 - 6·8%.^5,6^ While overall seroprevalence estimates of a given study depend on sampling method, assay characteristics, geography, and temporal factors, seroprevalence studies can provide information on the spread of COVID-19 that is missed by looking at the number of confirmed acute cases alone.

Seroprevalence studies are also useful for identifying demographic factors such as racial, ethnic and socioeconomic disparities among those exposed to SARS-CoV-2.^4,7,8^ The COVID-19 pandemic has been shaped by the deep and historic impacts of structural racism on disease disparities in US society as identified by serologic studies as well as hospitalization and mortality rates.^9,10^ For example, COVID-19 case and hospitalization rates among Black, Hispanic and Native American populations in the US, according to the Centers for Disease Control and Prevention, are 2.5-4.5 times higher than those in white populations^11^. Structural and occupational factors previously identified as drivers of race and ethnic disparities in health include unequal labor market opportunities and higher representation in essential work positions that lack job security, access to infection prevention control, benefits, and sick leave.^12–17^ Here, we confirm the findings of disparate SARS-CoV-2 exposure among racial and ethnic groups in the US by measuring seroprevalence in a large southern US health-care seeking cohort using remnant blood samples.

The following results are from the first six months (April 21-October 3, 2020) of an ongoing seroprevalence study using convenience remnant samples from clinical laboratories in central NC. The study catchment area covers Wake, Orange, Chatham, Johnston, Durham and Alamance counties and includes the county of the first confirmed case in NC,^18^ which occurred on March 3^rd^, 2020. On October 3^rd^, 2020, the cumulative total PCR and antigen-confirmed SARS-CoV-2 cases in the study catchment area was 52,722 (2·7% of the population), with 1,266 confirmed deaths.^19,20^ We used an in-house enzyme-linked immunoassay (ELISA) against the receptor-binding domain (RBD) of the spike protein of SARS-CoV-2^21^ and applied Bayesian inference^22^ to estimate seroprevalence and demographic risk factors of SARS-CoV-2 infection in a healthcare-seeking cohort over a six-month period.

## METHODS

### Sampling Strategy and Data Collection

Remnant plasma and serum samples were collected from four hospital-based clinical laboratories affiliated with the University of North Carolina (UNC) Health system. These laboratories receive and process clinical samples from inpatient units as well as outpatient clinics in NC. Each week, up to 300 remnant samples belonging to individuals 5-99 years of age were arbitrarily selected by the clinical laboratory for testing from each location. Samples were collected between April 21^st^, 2020 – October 3^rd^, 2020. Medical record numbers were recorded for each sample and duplicates were discarded. We abstracted the following demographic and clinical data from electronic medical records (EMR, Epic): age, sex, ethnicity, race, address including city, state and ZIP code, insurance coverage, insurance type, inpatient or outpatient status, encounter diagnosis (ICD-10 code), inpatient date of discharge, and whether or not COVID-19 testing was performed within a 30-day window prior to study sample collection. Written informed consent was not required due to the use of routinely collected samples. All data for this study were collected under UNC IRB #20-0791, which is conducted under Good Clinical Research Practices (GCP) and compliant with institutional IRB oversight. De-identified samples used for assay validation were collected under UNC IRBs #20-0913 and #08-0895.

### Enzyme-Linked Immunosorbent Assays

A total Ig and IgM SARS-CoV-2 RBD ELISA that does not react with common endemic human coronaviruses was used in this study as previously described.^21^ The spike protein N-terminal domain (NTD) antigen (16–305 amino acids, Accession: P0DTC2.1) was cloned into the pαH mammalian expression vector and purified using nickel-nitrilotriacetic acid agarose in the same manner. Each measurement was conducted in duplicate and duplicate values with variance > 25% and/or one value above assay cutoff were repeated. A correlation plot shows using 140 COVID-19 PCR-confirmed cases between our RBD Ig P/N ratios and the neutralization assay described below (**Figure S1**).

### Nucleocapsid protein ELISA

Detection of IgG antibody to SARS-CoV-2 N antigen was performed with the EUA approved Abbott SARS-CoV-2 IgG assay (Abbott Laboratories) on the Abbott Architect i2000SR immunoassay analyzer as previously described.^23^

### SARS-CoV-2 Neutralization Assays

To further characterize the SARS-CoV-2 antibody responses of this study, viral neutralization assays were obtained for 110 ELISA-positive samples that were selected randomly using the sample_n() function of the dplyr R package. Luciferase-expressing, full-length SARS-CoV-2 isolate WA1 strain (GenBank Accession#: MT020880) was engineered and recovered via reverse genetics and used to titer serially diluted sera on Vero E6 USAMRID cell as described previously. ^24^ The sample dilution at which a 50% reduction in RLU was observed relative to that of the virus control wells was used as the 50% neutralization titer (NT_50_) for that sample.

### Statistical Methods and Analyses

To account for plate-to-plate variability, we used positive to negative (P/N) ratios defined as the average optical density (OD) of the sample divided by the average OD of the negative control in the respective ELISA plate. Following the CDC recommendation to set specificity to 99 ·5%, we chose the 0.995 quantile of the P/N ratio for the negative validation samples as the P/N cutoff.^25^

We fit two statistical models to estimate seroprevalence. First, we fit a Bayesian autoregressive logistic model to estimate weekly prevalence across the six-month study period while accounting for uncertainty in the assay specificity and sensitivity due to finite lab validation samples. Second, we fit a Bayesian logistic regression model to estimate prevalence and conditional odds ratios by subpopulation with main effects for sex, race/ethnicity, age, in/out-patient status, and health insurance payor, while again accounting for uncertainty in the assay test characteristics (**Table S1**). Each group was compared to females, non-Latinx white, ages 5-17, outpatient, and private payor health insurance status as respective baseline categories. Details are given in Supplementary Methods: Bayesian seroprevalence models with unknown sensitivity and specificity. These Bayesian hierarchical models (BHM) simultaneously model study data and validation data to produce prevalence estimates and credible intervals that reflect both uncertainty due to the finite study sample as well as the uncertainty in the sensitivity and specificity of the ELISA, with statistical uncertainty represented by 95% credible intervals.

## RESULTS

### Cohort Characteristics

From April 21, 2020 – October 3, 2020, after excluding duplicate samples, 9,624 remnant samples were analyzed from four UNC Health hospitals in central North Carolina. The six counties most heavily sampled were Orange, Johnson, Chatham, Wake, Durham and Alamance, with 6,946 (72·2%) of individuals residing in these counties **(Figure 1)**. The study consists of 5,417 females (56·3%) and 4,206 males (43·7%) which is similar to the demographics of this region (**Table 1**). Less than 6% of individuals were in the youngest age group (5-17 years old), though this age group represents over 18% of the study area’s population. Approximately 90% of study individuals were insured, with 8% falling into the self-pay category. The majority of sampled individuals were seen at UNC Memorial Hospital, ∼3% were acute or trauma cases and ∼5% had a visit diagnosis of fever or respiratory symptoms (**Table S2**). Overall, approximately 1% of patients had an associated COVID-19 visit diagnosis, with a significant difference between inpatients (2 ·8%) and outpatients (0·3%) (Chi-squared test; p<0·0001) **(Table S3)**.

**Table 1.** Study participants by demographic factors of interest. Note, because of how the NC census reports data, the sex and age breakdowns of the 6-county demographics includes only individuals over the age of 4 (including those over age 99), but the race/ethnicity breakdown includes individuals of all ages. Additionally, the 65-99 age category is actually age 65+ for the 6-county demographics.

| Table 1. Study participants by demographic factors of interest. |  |  |  |  |  |  |  |
| --- | --- | --- | --- | --- | --- | --- | --- |
|  | 4/19-6/13 |  | 6/14-8/08 |  | 8/09-10/03 |  | 6-county |
|  | N | (%) | N | (%) | N | (%) | Demographics (%) |
| **Sex** |  |  |  |  |  |  |  |
| Female | 1947 | 56.2 | 2020 | 57.3 | 1450 | 55.1 | 51.8 |
| Male | 1515 | 43.7 | 1508 | 42.7 | 1183 | 44.9 | 48.2 |
| Unreported | 1 | 0.0 | 0 | 0.0 | 0 | 0.0 | — |
| **Age** |  |  |  |  |  |  |  |
| 5-17 | 259 | 7.5 | 163 | 4.6 | 150 | 5.7 | 18.4 |
| 18-49 | 1311 | 37.9 | 1052 | 29.8 | 830 | 31.5 | 48.7 |
| 50-64 | 926 | 26.7 | 1030 | 29.2 | 725 | 27.5 | 19.7 |
| 65-99 | 967 | 27.9 | 1283 | 36.4 | 928 | 35.2 | 13.1 |
| **Race/Ethnicity** |  |  |  |  |  |  |  |
| NL White | 2113 | 61.0 | 2267 | 64.3 | 1628 | 61.8 | 59.7 |
| NL Black | 845 | 24.4 | 803 | 22.8 | 603 | 22.9 | 21.0 |
| NL Other | 210 | 6.1 | 195 | 5.5 | 194 | 7.4 | 8.2 |
| Latinx | 295 | 8.5 | 263 | 7.5 | 208 | 7.9 | 11.1 |
| **In/Out patient** |  |  |  |  |  |  |  |
| Inpatient | 1057 | 30.5 | 961 | 27.2 | 839 | 31.9 | — |
| Outpatient | 2394 | 69.1 | 2562 | 72.6 | 1792 | 68.1 | — |
| Unknown | 12 | 0.3 | 5 | 0.1 | 2 | 0.1 | — |
| **Payor** |  |  |  |  |  |  |  |
| Public | 1825 | 52.7 | 2050 | 58.1 | 1509 | 57.3 | — |
| Private | 1249 | 36.1 | 1172 | 33.2 | 920 | 34.9 | — |
| Self-Pay | 326 | 9.4 | 254 | 7.2 | 181 | 6.9 | — |
| Other/Unknown | 63 | 1.8 | 52 | 1.4 | 23 | 0.8 | — |

**Figure 1.**
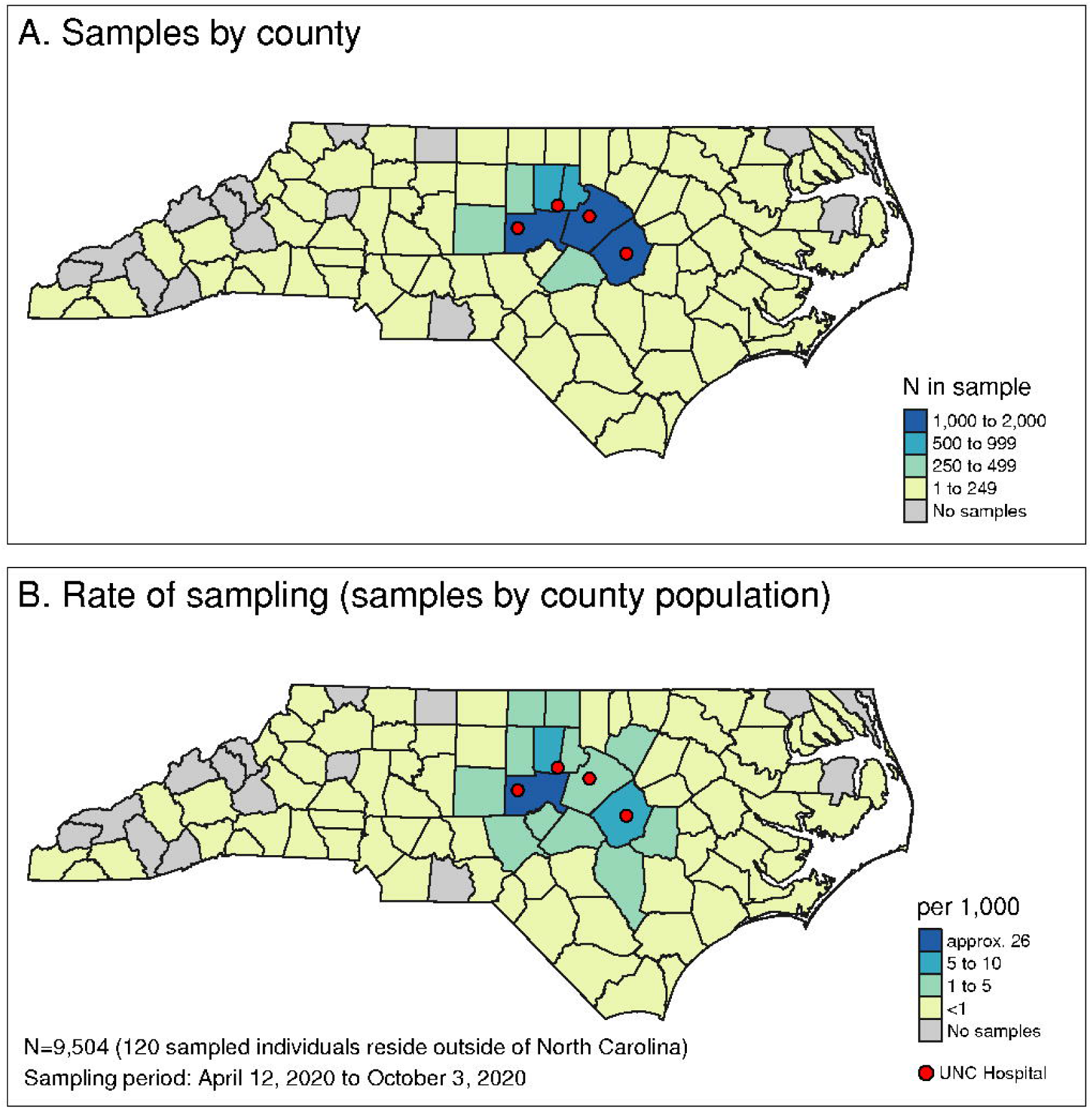
Catchment area for hospital remnant sample collection for UNC Health hospitals. Remnant samples were collected from hospital clinical laboratories from each of the four sites indicated by the red dots. (A) Number of samples collected by count as well as (B) the rate of sampling.^19^

### Overall seroprevalence estimates

The six-month period of the study was divided into three, two-month cohorts. The BHM-derived seroprevalence estimates increased from around 3% in April/May to around 9% in August/September **(Table 2**). Raw seroprevalence estimates also showed a similar increasing trend over the study period, but because they do not take into account assay performance uncertainty, they are slightly higher at ∼5% and ∼11%. Furthermore, seroprevalence estimates peaked in early August following a hospitalization peak in mid-July (**Figure 2A, 2C**). Cumulative PCR-positive COVID-19 cases reported by the state for these six counties increased over the study period (**Figure 2B**) with the most rapid accumulation of cases occurring from June to August. Unexpectedly, seroprevalence peaks followed by a slight decline, related to raw seroprevalence estimates at Johnston County hospital which surged from 7·81% in the first two months to 18·00% in the second two months coinciding with a peak in PCR-confirmed cases in the region, followed by a measured decline in raw seroprevalence to 14·80% in the final two-month period (**Table S6)**. This peak and decline was not affected by the removal of cases with ICD-10 visit codes for “COVID-19” or those we identify as “respiratory disease” (data not shown).

**Table 2.**
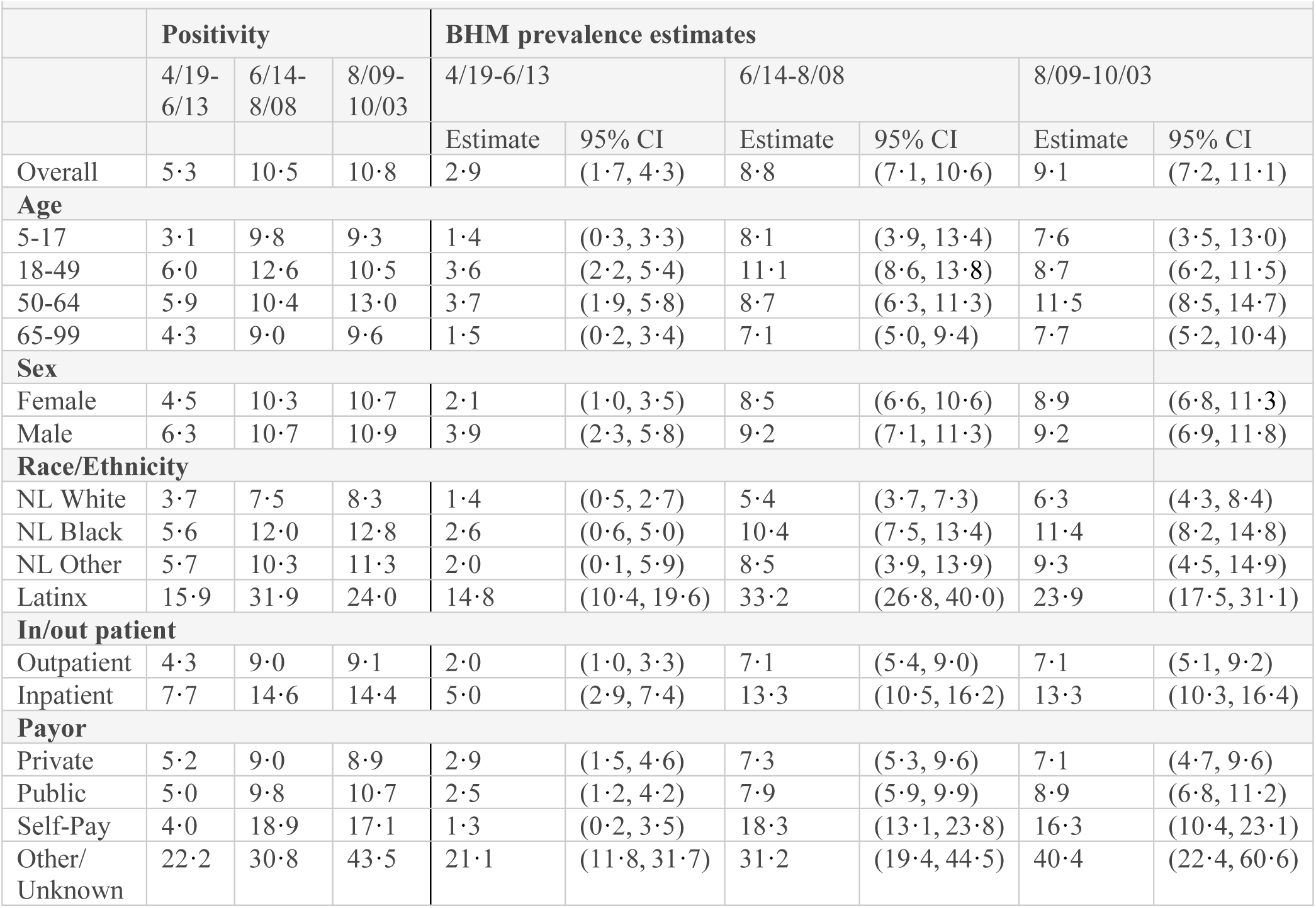
Cohort prevalence estimates. Raw seropositivity (%) and posterior mean seroprevalence estimates (%) from BHM with 95% credible intervals (lower bound, upper bound). NL, Non-Latinx.

**Figure 2.**
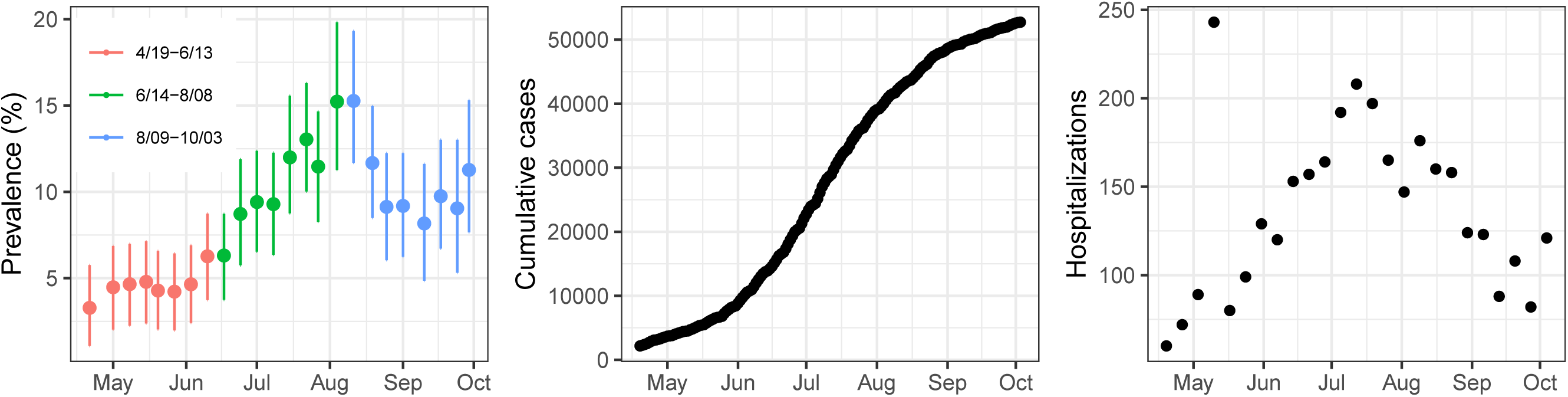
Trends in seroprevalence estimates. (A) Weekly posterior mean seroprevalence estimates and 95% credible intervals for the study period of 4/21-10/3 of the hospital samples by ELISA plotted over time over the course of the study period. (B) Cumulative daily COVID-19 PCR+ cases from the six-county area 4/19-10/3, and (C) weekly COVID-19 hospitalizations in the six-county area 4/19-10/3 from NC Department of Health and Human Services.

### Clinical and demographic differences in seroprevalence estimates

Latinx-identifying individuals have higher SARS-CoV-2 seroprevalence at 15-33% compared to non-Latinx individuals which have only 1-11% seroprevalence over the study period (**Table 2**). Individuals with “Other/Unknown” or “Self-pay” insurance status had a higher estimated seroprevalence (∼20-40% or ∼1-18%, respectively) than those with private or public health insurance (∼3-9%). Approximately 30% of Latinx individuals in this study were either in the other/unknown or self-pay health categories, disproportionately comprising ∼27% of these two categories but only accounting for ∼8% of our study population (**Table S5)**. To better compare the relative odds of SARS-CoV-2 seroprevalence for each clinical and/or demographic characteristic, we calculated conditional odds ratios for each variable we collected using the BHM (**Table 3**). Latinx individuals had the highest odds of SARS-CoV-2 exposure throughout the study period compared to non-Latinx white individuals, OR 7·77 overall (5·20, 12·10), ranging from 14·53 (6·47, 36·72) in the first two months to 4 ·34 (2·61, 7·41) in the last two months of the study. Individuals with unknown insurance status also had an elevated odds ratio of seropositivity at 3·81 (2·23, 6·54) compared to those with private insurance status. Over the entire period of the study, non-Latinx Black individuals, individuals aged 50-64 years, and inpatients, also had increased odds ratios of approximately two-fold compared to non-Latinx white individuals, individuals aged 0-17, and outpatients, respectively. The overall difference in odds ratios by age appears to be driven primarily by increased odds ratios in the first two months.

**Table 3.** Conditional odds ratios of being SARS-CoV-2 seropositive over the study period. Data is broken down into three two-month long periods in central North Carolina. Odds ratios of seropositivity calculated from the BHM with 95% credible intervals (lower bound, upper bound) are reported where the baseline groups for comparison are female, Non-Latinx white, age 5-17, outpatient, and private insurance. Odds ratios that do not overlap a value of one are bolded.

| **Table 3. Conditional odds ratios of being SARS-CoV-2 seropositive over the study period.** |  |  |  |  |  |  |  |  |
| --- | --- | --- | --- | --- | --- | --- | --- | --- |
|  | **4/19-6/13** |  | **6/14-8/08** |  | **8/09-10/03** |  | **4/19-10/03 (overall)** |  |
|  | Estimate | 95% CI | Estimate | 95% CI | Estimate | 95% CI | Estimate | 95% CI |
| **Sex** |  |  |  |  |  |  |  |  |
| Female | — | — | — | — | — | — | — | — |
| Male | **2.05** | **(1.08, 4.25)** | 1.10 | (0.80, 1.51) | 0.91 | (0.64, 1.29) | 1.27 | (0.98, 1.69) |
| **Race/Ethnicity** |  |  |  |  |  |  |  |  |
| NL White | — | — | — | — | — | — | — | — |
| NL Black | 1.66 | (0.53, 4.28) | **1.94** | **(1.31, 2.92)** | **1.82** | **(1.20, 2.79)** | **1.80** | **(1.19, 2.65)** |
| NL Other | 1.26 | (0.12, 5.74) | 1.58 | (0.74, 3.19) | 1.81 | (0.87, 3.57) | 1.54 | (0.66, 2.84) |
| Latinx | **14.53** | **(6.47, 36.72)** | **7.43** | **(4.70, 11.97)** | **4.34** | **(2.61, 7.41)** | **7.77** | **(5.20, 12.10)** |
| **Age** |  |  |  |  |  |  |  |  |
| 5-17 | — | — | — | — | — | — | — | — |
| 18-49 | 3.09 | (0.99, 11.43) | 1.38 | (0.68, 3.05) | 0.89 | (0.42, 2.03) | 1.56 | (0.92, 2.77) |
| 50-64 | **3.62** | **(1.13, 13.56)** | 1.34 | (0.64, 2.99) | 1.56 | (0.76, 3.54) | **1.96** | **(1.15, 3.55)** |
| 65-99 | 1.62 | (0.28, 6.90) | 1.49 | (0.71, 3.34) | 1.13 | (0.52, 2.64) | 1.40 | (0.71, 2.61) |
| **In/out patient** |  |  |  |  |  |  |  |  |
| Outpatient | — | — | — | — | — | — | — | — |
| Inpatient | **2.50** | **(1.31, 5.10)** | **1.91** | **(1.38, 2.68)** | **1.92** | **(1.34, 2.80)** | **2.09** | **(1.59, 2.85)** |
| **Payor** |  |  |  |  |  |  |  |  |
| Private | — | — | — | — | — | — | — | — |
| Public | 0.85 | (0.41, 1.73) | 0.89 | (0.58, 1.34) | 1.16 | (0.74, 1.85) | 0.96 | (0.70, 1.30) |
| Self-Pay | **0.18** | **(0.03, 0.64)** | **1.78** | **(1.07, 2.93)** | **1.94** | **(1.03, 3.63)** | 0.85 | (0.45, 1.41) |
| Other/<br>Unknown | **3.08** | **(1.15, 8.23)** | **2.73** | **(1.25, 5.98)** | **6.60** | **(2.29, 18.71)** | **3.81** | **(2.23, 6.54)** |

### SARS-CoV-2 RBD positive subset analysis

To determine the SARS-CoV-2 antibody repertoire in a subset of RBD Ig seropositive individuals, we randomly selected 110 participants and tested their sera for: RBD IgM, NTD IgG, and SARS-CoV-2 neutralizing antibodies. About 75% of individuals were positive for RBD IgM, 60% had NTD IgG antibodies, and about 50% had detectable neutralizing antibodies (**Figure 3A**). Of the participants with detectable functionally neutralizing antibodies, 23% had a high titer > 1:1280, 47% had a moderate titer of 1:160-1:1279, and 30% had a lower titer of 1:10-1:159. Furthermore, RBD Ig P/N antibody signal correlated more strongly with functionally neutralizing antibody levels (**Figure 3B**), than NTD IgG signal (**Figure 3C**). We also found that 36% (29/80) of those in this subset with an ICD-10 code binned as “Other” had detectable neutralizing antibodies, while 83% (25/30) of individuals with an ICD-10 code of “COVID-19” or what we identify as “respiratory disease” had neutralizing antibodies (**Figure 3D**). There was substantial agreement between the RBD Ig ELISA results reported here and 150 study individuals for which a clinical SARS-CoV-2 nucleocapsid IgG (Abbott assay) was available (Cohen’s kappa=0·685) **(Table S4)**.

**Figure 3.**
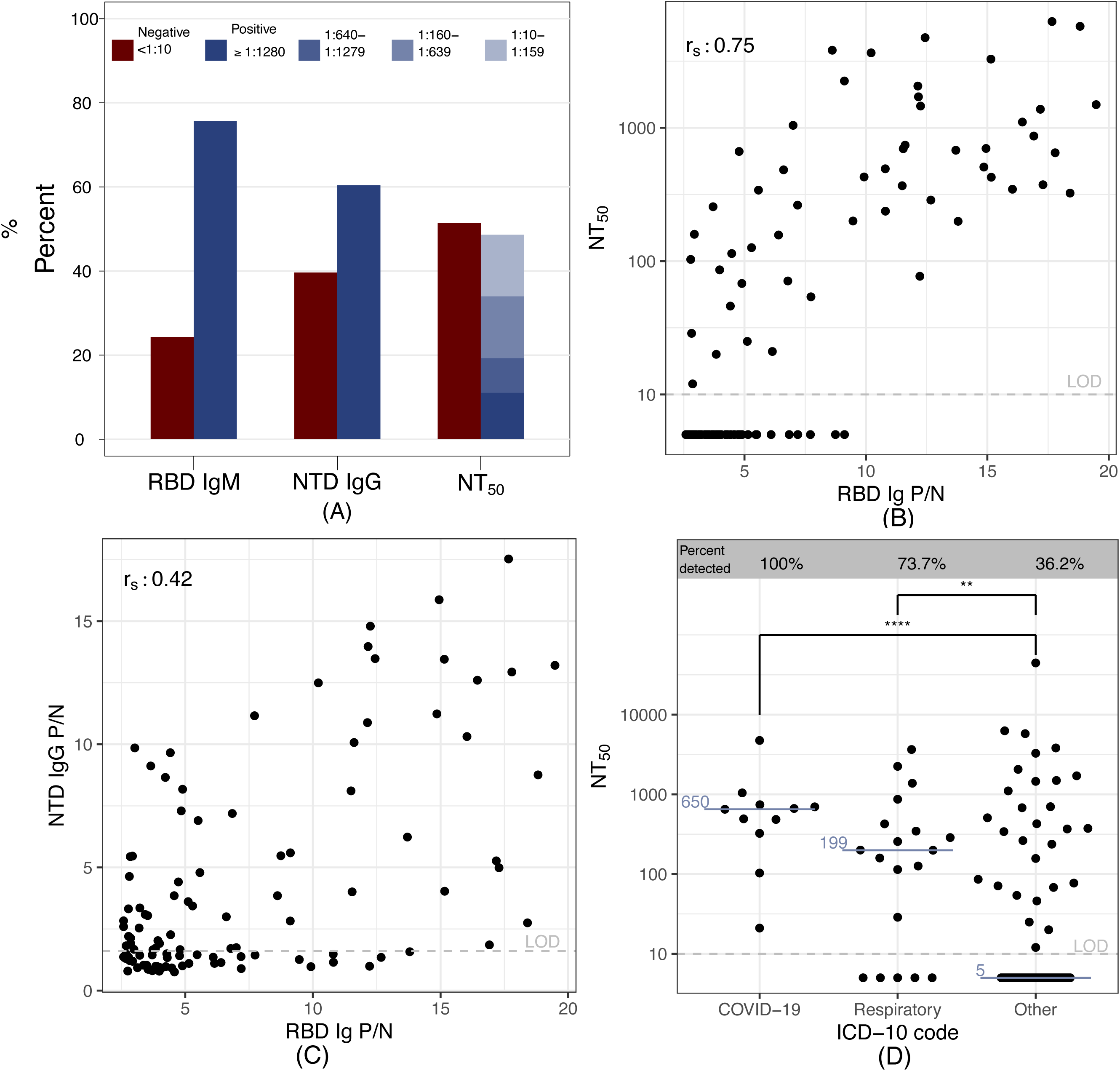
Antibody repertoires in an RBD Ig positive subset. 110 RBD Ig positive samples were chosen at random to undergo SARS-2 antibody repertoire analysis. (A) Percent of individuals with RBD IgM, NTD IgG and functionally neutralizing antibodies (NT50). (B) Correlation plot of NT50 and RBD Ig. (C) Correlation plot of NTD IgG and RBD Ig, r_s_ = Spearman correlation coefficient displayed in the top left of panels (B) and (C). (D) NT50 values for each diagnosis binning category based on ICD-10 codes. Medians shown in blue. Two-tailed Mann-Whitney, ****p<0.0001, **p=0.0078.

## DISCUSSION

Here we describe SARS-CoV-2 seroprevalence in a total of 9,624 unique healthcare-seeking individuals in central North Carolina using clinical remnant samples from four regional hospitals between April and October 2020. Employing a Bayesian framework^22^ to capture assay uncertainty in both field and lab validation data, we estimate a significant increase in overall seroprevalence from 2 ·9% (95% CI 1·7% - 4·3%) at the start of the study period, to 9·1% (95% CI 7·2% - 11·1%) at the end of the study period, approximately six months after the first case in the state.

The end-of-study prevalence identified here is significantly higher than the cumulative number of cases identified by PCR or antigen testing in the same county region at the same date, though determining the degree to which the identified cases undercount true infections requires more representative sampling.

A previous study from central North Carolina that overlaps with the first two months of our study period found seroprevalence in an asymptomatic healthcare-seeking cohort below 1% using the Abbott nucleocapsid IgG assay.^5^ This is much lower than the ∼3 % seropositive estimate in our cohort over this time period, and may be due to under-sampling of Latinx individuals in that study and/or preferential sampling of asymptomatic individual. There is also growing concern about the use and performance of nucleocapsid IgG assays in individuals with asymptomatic or mild disease.^26^ The nationwide CDC study that used remnant clinical samples from inpatients and outpatients found a seroprevalence of 6·8% in NC in September 2020, which is closer to our estimate of 9 ·1% during the final two months of this analysis.

The conditional odds ratios we calculated assume that all other variables are held constant while estimating the effect of one demographic variable at a time. We found that Latinx individuals had the highest odds of SARS-CoV-2 seropositivity, and that non-Latinx Black individuals also had high odds of SARS-CoV-2 seropositivity, corroborating previous observations.^4,7,8^ The high odds ratios by race and ethnicity decrease over time, consistent with the virus spreading first among individuals with high exposure risk and later to the rest of the population. Residential segregation, crowded households, socioeconomic disadvantage, mass incarceration, and inequities in access to insurance, health care, and access to testing, vaccination, and treatments have all been cited as factors that have contributed to the large and sustained racial and ethnic disparities in COVID-19 in the US.^13,15,27–29^ We also observed that individuals that fell into the “self-pay” category for their healthcare or otherwise had unknown healthcare status had higher SARS-CoV-2 seropositivity and odds ratios. The significant overlap in the Latinx population and these insurance categories is concerning because the high odds ratios and seroprevalence in these categories can lead to much higher exposure risk among the significant number of underinsured Latinx individuals^30^.

Studies of PCR-positive symptomatic COVID-19 cases have reported good neutralizing antibody responses in these individuals.^31^ Thus, it was surprising that we observed 51% of individuals in our RBD-positive subset analysis did not have detectable neutralizing antibodies. Though we do not know what proportion of individuals in our study had asymptomatic infections, low neutralizing antibody titers may be explained by short duration of viral replication in respiratory compartments and low to no viral replication in the serum or blood of those with mild or asymptomatic disease. Not surprisingly, when we looked at our neutralizing antibody results by ICD-10 code, the majority of all individuals with a “respiratory disease” or “COVID-19” diagnosis had developed neutralizing antibodies. Reports of mild disease COVID-19 cohorts support the idea that detectable neutralizing antibody titers are not necessarily identified after mild COVID-19.^23,32^ In this subset analysis we also found that 75% had RBD IgM antibodies, indicating that their infections likely occurred within the past three months.^23^ Furthermore, a majority of individuals in this subset had detectable NTD IgG antibodies; the NTD has recently been found to be an important target for the B.1.1.7, B.1.351, and B.1.1.28.1 SARS-CoV-2 variants.^33^

The primary limitation of this study is that the study population, composed of individuals accessing care at UNC area hospitals and clinics may differ from the overall population in central North Carolina in ways that are not captured in demographic data (e.g., overall health status). Accordingly, we have chosen to not weight our dataset to county demographics and therefore do not provide overall estimates of seroprevalence in the six-county area as that would require more representative sampling methodology.^34^ Furthermore, many clinics and hospital elective procedures were closed or only seeing patients virtually during the first few months of the study period.

The unexpected seroprevalence peak observed at the Johnston County hospital suggests that the population accessing care at these clinical sites did not have consistent exposure risk over time. As expected, seroprevalence estimates in this cohort track closely with COVID-19 hospitalizations in the four hospitals in this study with a two-week lag which could be due to time to seroconvert. Declining antibody over this time period to undetectable levels is unlikely, as the length of the study is shorter than it takes for significant antibody decline to undetectable levels, although little is known about antibody levels over time in the asymptomatic population.^31^

Other limitations of the study include that we could not break down odds ratios by all races and/or by race and ethnicity at the same time, or by multiracial categories because the number of individuals became too small to allow broad interpretation. Finally, though the “self-pay” insurance category includes the uninsured, we cannot confidently state that everyone in this category was uninsured because lack of insurance is not a specific category that is captured in the EMR. Although SARS-CoV-2 seroprevalence of healthcare-seeking individuals is an imperfect comparison to the general population, we maintain that it is a useful sentinel population to understand overall trends, especially when attempting to surveil rural populations residing in areas without strong public health systems and spread over a large geographic area.

Based on our estimates of seroprevalence in the population accessing healthcare, cumulative case numbers confirmed by molecular diagnostics are likely under-representing the true number of cases. Public health distancing measures, mask wearing, and vaccination should continue to be prioritized in order to lower the transmission of SARS-CoV-2 and subsequent loss of lives. Our findings of a significantly higher odds of SARS CoV-2 seropositivity among Latinx and non-Latinx Black populations corroborate numerous studies describing large racial and ethnic disparities in SARS-CoV-2 infection, morbidity and mortality in the US. ^4,7,8^ Vaccination programs should address structural and occupational factors that drive race and ethnic disparities in health outcomes in the US to ensure that individuals at particularly high exposure risk of SARS-CoV-2 have timely access to SARS-CoV-2 vaccination.

## Data Availability

Data Sharing
Deidentified individual data will be shared beginning 9 to 36 months following publication provided the investigator who proposes to use the data has approval from an Institutional Review Board (IRB), Independent Ethics Committee (IEC), or Research Ethics Board (REB), as applicable, and executes a data use/sharing agreement with UNC.

## Acknowledgements

We would like to thank Jennifer Dan, Alessandro Sette and Shane Crotty’s Laboratories at La Jolla Institute of Immunology for supplying some of the serum controls in our validation cohort.

## Data Sharing

Deidentified individual data will be shared beginning 9 to 36 months following publication provided the investigator who proposes to use the data has approval from an Institutional Review Board (IRB), Independent Ethics Committee (IEC), or Research Ethics Board (REB), as applicable, and executes a data use/sharing agreement with UNC.

## Author contributions

CL, CHC: Conceptualization, Data curation, Investigation, Project Administration, Writing – original draft. SP, KB: Formal analysis, Software, Visualization. MD, QG, UPV: Data curation, Investigation, Project Administration. SJG: Formal analysis, Software, Visualization. YJH: Investigation. PL: Methodology, Resources. RR, MG, CW, KP, CA: Resources. JS: Conceptualization, Resources. ME: Conceptualization, Formal analysis, Resources, Supervision. RB, AA: Conceptualization, Funding acquisition. BKF: Conceptualization, Formal analysis, Resources, Software, Supervision. DBL: Conceptualization, Formal analysis, Software. ADS: Conceptualization, Funding acquisition, Methodology, Project Administration, Resources, Supervision. JJJ: Conceptualization, Funding acquisition, Project Administration, Supervision. AJM: Conceptualization, Funding acquisition, Investigation, Project Administration, Supervision, Writing – original draft. CL, CHC, SP, KB, MD, QG, UPV, SJG, YJH, PL, JS, ME, RB, AA, BKF, DBL, ADS, JJJ, AJM: Writing- reviewing & editing.

