## Supplemental Appendix for "Disparities in SARS-CoV-2 seroprevalence among individuals presenting for care in central North Carolina over a six-month period"

### Supplementary Appendix

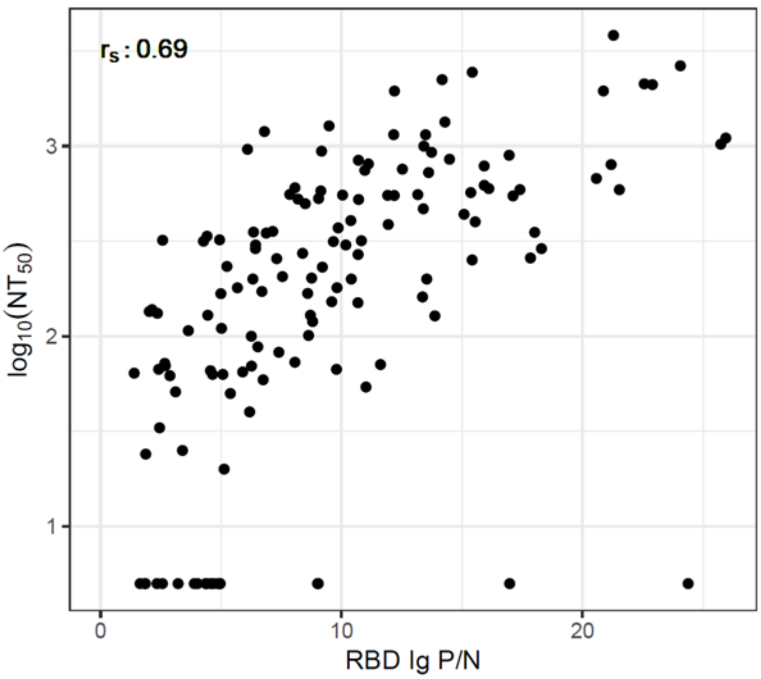

**Figure S1. Correlation plot between RBD Ig P/N and neutralization assay.** Spearman correlation coefficient was obtained ($r_s$), $p < 0{\cdot}0001$.

| Table S1. ELISA Validation Data | | |
|---|---|---|
| | **% Sensitivity (95% CI)** | **% Specificity (95% CI)** |
| RBD total Ig ELISA (≥ 9 days post symptom onset) | 89·7% (130/145) (84·7, 94·6) | 99·3% (272/274) (98·3, 100·0) |
| PCR+ controls (n = 145) | N = 32 (Crotty Lab, La Jolla) | |
| | N = 113 (UNC CP donor cohort) | |
| Negative controls (n = 274) | N = 122 (UNC pre-2019 healthy adults) | |
| | N = 48 (UNC pre-COVID-19 arboviral samples, TB endemic region) | |
| | N = 44 (UNC, clinical pre-organ transplant) | |
| | N = 28 (UNC, clinical HIV+) | |
| | N = 16 (healthy adults, Crotty Lab, La Jolla) | |
| | N = 16 (UNC, respiratory illness samples, COVID-19 negative) | |

**Table S1. ELISA Validation Data.** CI, confidence interval; PCR, polymerase chain reaction; CP, convalescent plasma; TB, tuberculosis.

**Table S2. Study Individual Numbers by Clinical Factors.**

| | 4/19-6/13 | | 6/14-8/08 | | 8/09-10/03 | |
|---|---|---|---|---|---|---|
| | N | (%) | N | (%) | N | (%) |
| **Hospital** | | | | | | |
| Chatham Hospital | 604 | 17·4 | 909 | 25·8 | 298 | 11·3 |
| UNC Hospitals | 1490 | 43·0 | 1491 | 42·3 | 1291 | 49·0 |
| Johnston Hospital | 627 | 18·1 | 639 | 18·1 | 473 | 18·0 |
| Rex Hospital | 742 | 21·4 | 489 | 13·9 | 571 | 21·7 |
| **In/Outpatient** | | | | | | |
| Inpatient | 1057 | 30·5 | 961 | 27·2 | 839 | 31·9 |
| Outpatient | 2394 | 69·1 | 2562 | 72·6 | 1792 | 68·1 |
| Unknown | 12 | 0·3 | 5 | 0·1 | 2 | 0·1 |
| **Visit type** | | | | | | |
| Traumatic | 104 | 3·0 | 103 | 2·9 | 62 | 2·4 |
| Not traumatic | 2961 | 85·5 | 3066 | 86·9 | 2180 | 82·8 |
| Unknown | 398 | 11·5 | 359 | 10·2 | 391 | 14·8 |
| **Condition** | | | | | | |
| Respiratory | 173 | 5·0 | 167 | 4·7 | 75 | 2·8 |
| COVID-19 | 31 | 0·9 | 35 | 1·0 | 30 | 1·1 |
| Other | 2861 | 82·6 | 2967 | 84·1 | 2137 | 81·2 |
| Unknown | 398 | 11·5 | 359 | 10·2 | 391 | 14·8 |
| **Payor** | | | | | | |
| Public | 1825 | 52·7 | 2050 | 58·1 | 1509 | 57·3 |
| Private | 1249 | 36·1 | 1172 | 33·2 | 920 | 34·9 |
| Self-Pay | 326 | 9·4 | 254 | 7·2 | 181 | 6·9 |
| Other/Unknown | 63 | 1·8 | 52 | 1·4 | 23 | 0·8 |

**Table S2. Study Individual Numbers by Clinical Factors.**

| Table S3. Rates of COVID-19 Visit Codes for Inpatients and Outpatients. | | |
|---|---|---|
| | **Inpatient** | **Outpatient** |
| **COVID-19 Visit Code** | 80/2828 (2·8%) | 16/5646 (0·3%) |

**Table S3. Rates of COVID-19 Visit Codes for Inpatients and Outpatients.**

| Table S4. Subset of individuals with recorded Abbott IgG | | |
|---|---|---|
| | **Abbott Nucleocapsid ELISA negative** | **Abbott Nucleocapsid ELISA positive** |
| **RBD Ig ELISA negative** | 132 | 3 |
| **RBD Ig ELISA positive** | 5 | 10 |

**Table S4. Subset of individuals with recorded Abbott IgG.** RBD Ig ELISA results for 150 patients who received a UNC hospital lab-based Abbott nucleocapsid IgG ELISA within one month prior to study enrollment.

| S5. Insurance category by race/ethnicity | | | | | | | | |
|---|---|---|---|---|---|---|---|---|
| | **Private** | | **Public** | | **Self-Pay** | | **Other/Unknown** | |
| | N | (%) | N | (%) | N | (%) | N | (%) |
| **NL White** | 2173 | 36·17 | 3439 | 57·24 | 347 | 5·78 | 49 | 0·82 |
| **NL Black** | 641 | 28·48 | 1402 | 62·28 | 177 | 7·86 | 31 | 1·38 |
| **NL Other** | 323 | 53·92 | 223 | 37·23 | 42 | 7·01 | 11 | 1·84 |
| **Latinx** | 204 | 26·63 | 320 | 41·78 | 195 | 25·46 | 47 | 6·14 |
| **TOTAL** | 3341 | 34·72 | 5384 | 55·94 | 761 | 7·91 | 138 | 1·43 |

**Table S5. Count and percentage in each insurance category by race/ethnicity.**

| Table S6. Raw sample positivity by hospital | | | |
|---|---|---|---|
| | **4/19-6/13** | **6/14-8/08** | **8/09-10/03** |
| **Johnston Hospital** | 7·81 | 18·00 | 14·80 |
| **Chatham Hospital** | 7·45 | 8·69 | 12·08 |
| **UNC Hospitals** | 4·03 | 9·93 | 10·38 |
| **Rex Hospital** | 4·04 | 5·93 | 7·71 |

**Table S6. Raw antibody test positivity (percent) by hospital.**

**Supplementary Methods: Bayesian seroprevalence models with unknown sensitivity and specificity**

#### 1 Determining Test Results

##### 1.1 Quantitative test outcome

To account for plate-to-plate variability (i.e., batch effects), similar to Zhang and colleagues,[1] we used P/N ratios, rather than using the raw optical density (OD) values, defined as

$$\text{P/N}=\frac{average\ OD\ sample}{average\ OD\ negative\ control}$$

where the negative control values were those from the same plate as the sample. Accounting for batch effects in the P/N ratio removes the need for defining plate specific cutoffs, and rather we can define one cutoff on how many times larger the sample's OD value is relative to the corresponding negative control's OD value.

##### 1.2 Cutoff Selection

The CDC recommends selecting a threshold such that the test has 99·5% specificity.[2] We followed this recommendation here specifying the cutoff to be the standard estimate of the 0·995 quantile (based on the quantile function in R) of the negative lab samples. Using the 274 negative controls, the cutoff was 2·57 with empirical sensitivity of 89·7% and empirical specificity of 99·3%. Therefore, a sample is considered positive if its average OD value is 2·57 or more times larger than the average OD of the corresponding plate negative controls.

#### 2. Temporal Logistic Model

We fit a Bayesian autoregressive logistic model to estimate weekly prevalence while accounting for uncertainty in test sensitivity and specificity. Let $n_t$ give the number of samples in week t, and $y_t$ give the number of samples that tested positive in week t, for $t \in \{1,...,T = 24\}$. Then

$$y_t \sim \text{binomial}(n_t,p_t) \quad t = 1,...,T$$

were $p_t$ gives the probability of a positive test in week t. To account for the error rate of the test, we define

$$p_t = \pi_t \text{sens}+(1-\pi_t)(1-\text{spec})$$

where $\pi_t$ is the probability an individual has COVID-19 antibodies in week t, sens gives the sensitivity of the test, and spec gives the specificity of the test.

Assuming seroprevalence varies smoothly, we define an AR(1) process for the $\pi_t$ as follows. First, let $\beta_t = \text{logit}(\pi_t)$. Then we model $\beta_t$ as

$$\beta_t \sim \text{normal}(\alpha+\varphi\beta_{t-1},\sigma_\beta^2) \quad t=2,...,T$$

$$\beta_1 \sim \text{normal}(\alpha, 0{\cdot}5).$$

As we expect autocorrelation and we are on the logit scale, we expect $\sigma_\beta^2$ to be relatively small, so a relatively vague prior is assumed

$$\sigma_\beta^2 \sim \text{normal}^+(0,0{\cdot}5),$$

where $\text{normal}^+$ indicates the folded normal distribution. We found changing the prior variance of $\sigma_\beta^2$ had minimal effect on the estimates and associated uncertainty of $\{\pi_t\}$. Similarly, we put vague priors on $\alpha$, $\varphi$, sens, and spec:

$$\alpha \sim \text{logistic}(0, 1);$$

$$\varphi \sim \text{normal}(0, 1);$$

$$\text{sens} \sim \text{uniform}(0, 1);$$

$$\text{spec} \sim \text{uniform}(0, 1).$$

Finally, to estimate sensitivity and specificity, we assume

$$y_{spec} \sim \text{binomial}(n_{spec}, \text{spec}),$$

$$y_{sens} \sim \text{binomial}(n_{sens}, \text{sens})$$

where $y_{spec}$ is the number of negative controls that tested negative out of $n_{spec}$ negative controls. Similarly, $y_{sens}$ is the number of positive controls that tested positive out of $n_{sens}$ positive controls.

##### 3. Logistic Regression Model

We fit a Bayesian logistic regression model with main effects for sex, race/ethnicity, age, in/out-patient status, and payor. Interactions were considered, but not found to significantly improve the fit. This model allows us to simultaneously model the hospital data and the lab validation data.

To ensure each category in our main effects had a sufficient sample size, some categories were collapsed. All outpatient, emergency, or unknown patients were listed as "outpatient." Additionally, the "other" and "unknown" categories for payor were collapsed. Finally, the one patient with sex listed as "X" was removed from the dataset for this analysis.

We define the likelihood

$$y_i \sim \text{Bernoulli}(q_i) \; i = 1,...n$$

where $y_i$ is an indicator for whether individual i tests positive for COVID-19 antibodies and $q_i$ is the probability of a positive test for individual i. The number of patients is given by n. To account for the error rate of the test, similar to the temporal model, we define

$$q_i = \pi_i \text{sens} + (1-\pi_i)(1-\text{spec})$$

where $\pi_i$ is the probability individual i has COVID-19 antibodies, sens gives the sensitivity of the test, and spec gives the specificity of the test. Finally, the probability a patient has COVID-19 antibodies is assumed to equal

$$\pi_i = \text{logit}^{-1}\,(\beta_{0t_i} + \mathbf{x}'_i \boldsymbol{\alpha}_{t_i})$$

for the vector of p predictors $\mathbf{x}_i$, coefficients $\boldsymbol{\alpha}_t$, and intercept $\beta_{0t}$, where $t_i$ gives the time period patient i was sampled during, $t \in \{1, \ldots, T = 3\}$. This allows the intercept and coefficients to vary across time periods, but the sensitivity and specificity estimates to be pooled across time. For this analysis, the vector $\mathbf{x}$ contains indicators for Male, NL Black, NL Other, Latinx, age 18-49, age 50-64, age 65-99, outpatient, public payor, self-pay, and unknown payor. This leaves inpatient, private paying, NL White, females aged 5-17 as the baseline category. Let $\mathbf{x}_i$ be the $i^{th}$ row of the n×p matrix $\mathbf{X}$. We calculated the effect of the covariates over the entire study period as

$$\bar{\boldsymbol{\alpha}} = \sum_{t=1}^{T} \boldsymbol{\alpha}_t / T,$$

and present $exp(\bar{\boldsymbol{\alpha}})$ as the average estimated odds ratio.

As before, to estimate sensitivity and specificity, we assume

$$y_{spec} \sim \text{binomial}(n_{spec}, \text{spec})$$

$$y_{sens} \sim \text{binomial}(n_{sens}, \text{sens})$$

where $y_{spec}$ is the number of negative controls that tested negative out of $n_{spec}$ negative controls and $y_{sens}$ is the number of positive controls that tested positive out of $n_{sens}$ positive controls.

We chose non-informative priors:

$$\boldsymbol{\alpha}_t \sim \text{normal}(\mathbf{0}_p, 2\mathbf{I}_p)$$

$$\text{sens} \sim \text{uniform}(0, 1)$$
$$\text{spec} \sim \text{uniform}(0, 1)$$

$$\beta_{0t} + \bar{\boldsymbol{x}}_t'\boldsymbol{\alpha}_t \sim \text{logistic}(0,1) \quad t = 1, \dots, T$$

where $\bar{\boldsymbol{x}}_t$ is the p-dimensional vector of the average of each column of $\mathbf{X}$ across the rows corresponding with time period t (i.e. the proportion of observations in each group during that time period). In this way, following Gelman and Carpenter (2020)[3], the prior on $\beta_0$ models the probability of the average patient being seropositive as uniform on the interval (0,1). Note, because the $\boldsymbol{\alpha}_t$ coefficients are on the logit scale, a variance of 2 is relatively vague. For example, this places about 68% probability that an element of $\boldsymbol{\alpha}_t$ is between -√2 and √2, evaluating to a subpopulation having 0·24 to 4·11 times the seroprevalence of another.

An artifact of the model accounting for uncertainty in test sensitivity and specificity is that when there is low observed positivity, small changes in the estimated specificity can result in large changes in the overall seroprevalence, compared to when there is larger overall positivity. Therefore, there is more uncertainty in these cases (as we observed in earliest time period of our data compared to the latter two). This is because when there are very few positive tests, small declines in the estimated specificity suggest the observed positives should be classified as false positives and the seroprevalence is very low. Without copious amounts of lab validation data, some uncertainty in specificity is expected and this uncertainty will propagate to the seroprevalence and coefficient estimates.

**4. MCMC Algorithm**

The models were fit using a Markov chain Monte Carlo algorithm implemented in Stan.[4] For each model, we ran four chains 5000 iterations each with the first 2500 iterations used as burn-in. The $\hat{R}$ value[5] was 1 for each parameter, suggesting convergence. The effective sample size was over 2300 for each parameter in the logistic regression model, and over 1000 for each parameter in the temporal model.

**5. Quantifying Uncertainty**

For results from the Bayesian models, we reported posterior means and equal-tail 95% credible intervals (i.e., the 2.5% and 97.5% quantiles of the posterior draws). In Table S1, we calculated standard 95% confidence intervals:

$$\hat{p} \pm Z_{\alpha/2}\sqrt{\frac{\hat{p}(1-\hat{p})}{n}}$$

where $\hat{p}$ denotes the observed proportion, $n$ is the sample size, and $Z_{\alpha/2}$ is the $\alpha/2$ quantile of the standard normal distribution.

**6. Demographic data categorization**

To categorize individual clinical encounters associated with the blood draws we sampled, we obtained ICD-10 codes from any inpatient or outpatient visit at the same location within fourteen days of when we received and sampled the blood draw. We prioritized inpatient visits over outpatient visits unless no inpatient visit was available. If there was no visit within the past fourteen days of the blood draw, we instead used the visit closest to the most recent specimen collection date within a thirty-day period. Individuals with no visit at the same location within thirty days of their blood draw were excluded from analysis. To capture any upper respiratory infection, respiratory disease due to external agents, interstitial lung disease, imaging abnormalities of the lung, cough, fever, and dyspnea, we used the International Classification of Diseases, 10th revision (ICD-10) codes J00-J006, J009-J018, J20-J22, J40-J47, J60-J70, J80-J84, J96-J99, R91, R05, R06.0, and R50. COVID-19 diagnosis was defined as presence of the U07·1 code in the visit nearest the sampled blood draw. Likewise, acute or trauma cases were defined as any of the following ICD-10 codes: O00, O01, O02, O03, O04, O07, O08, O015·1, all S codes, all T codes (except T36-T39, T41, T46, T50, T80-T88), all V codes, all W codes, all X codes, all Y codes (except Y62-Y84 and Y-90-99).

Insurance status was determined from the most recent clinical encounter prior to the sampled blood draw. “Private Insurance” was classified as any of the following listed for a patient’s visit: Blue Cross/Blue Shield, Private health insurance, or State Government insurance. “Public Insurance” was classified as any of these following: Medicaid applicant, Medicaid, Medicare, Department of Veteran’s Affairs, Tricare, and Corrections State insurance. “Self-pay” includes anyone paying out of pocket. “Unknown/Other” consists of individuals for whom the health insurance payor was left blank or otherwise unidentifiable, as well as listed insurance that read “Legal Liability / Liability Insurance”, “Other specified but not otherwise classifiable (includes Hospice - Unspecified plan)”, and “Other”.

Race and ethnicity identity was ascertained from that listed in the EMR for each patient. The categories listed under Epic’s EMR that we received included “American Indian or Alaska Native”, “Asian”, “Black or African American”, “Native Hawaiian or other Pacific Islander”, “Other Race”, “Patient Refused”, “Unknown” or “White or Caucasian”. For ethnicity, we received information on whether patients self-identified as “Hispanic or Latino”, or were listed as “Patient Refused” or “Unknown”. In our report, we collapse race and ethnicity from separate variables into a single variable in order to investigate the impact of systemic racism on SARS-COV-2 seroprevalence by both race and ethnicity at the same time, though the constructs of race and ethnicity are inherently surrogate measures of racism and other forms of marginalization.[6]

We therefore binned individuals into the following groups: “Black or African American” that indicated “Non-Hispanic or Latino,” “Patient Refused,” or “Unknown” were binned as “Non-Latinx Black”, similarly for “White or Caucasian” as “Non-Latinx White”, similarly for all other groups as “Non-Latinx Other”. Anyone that indicated “Hispanic or Latino” were binned as “Latinx”, and therefore could self-identify as any of the above race categories. We do not further separate out other intersections of race and ethnicity because the number of individuals becomes too small to make conclusive claims on odds of seropositivity. We here opt to use Latinx in place of “Hispanic” though it is not the only way to refer to this grouping of individuals that often share cultural characteristics, language, religion, and ancestral geography and history.[7] We also compare racial, ethnic, and age demographics in the study population to the demographics of the 6-county area where most of the study population resided using data collected from US Census Data.[8]
